# From protection of sacrificial self to critical turning points and growth: Nurses’ experiences on the frontline during the COVID-19 pandemic

**DOI:** 10.1101/2024.11.19.24317599

**Authors:** Sinéad Creedon, Anna Trace

## Abstract

The mental health and wellbeing of nurses has been a concern, long before the arrival of the COVID-19 pandemic. Working on the frontline under such challenging circumstances, for extended periods of time, has resulted in negative psychological responses. The current study aims to examine nurses’ resilience in acute hospitals in Ireland, during a period of adversity (pandemic), to explore it’s impact on their personal and professional identity, and their perception of meaningful supports and coping. An interpretative phenomenological analysis was carried out to gain insight into how nurses adapted to the changing work environment during the pandemic. Online semi-structured interviews were carried out with six experienced female nurses, who were redeployed to the frontline from their own roles. Three experiential themes representing the nurses’ journey were generated: Protection of Sacrificial Self; The Fortifying Effect of Us, and Critical Turning Points & Growth. Nurses made significant sacrifices and had to find ways to detach to cope. They revitalized themselves by creating a sense of ‘us’ to help them face a harsh climate against others, which enabled critical turning points and growth. This study has strongly highlighted the emotional effects on nurses due to feeling isolated, undervalued, and excluded during redeployment to the frontline. It has also featured how well nurses coped while faced with an existential crisis and has given voice to all nurses who faced this pandemic despite exposure to risk of burnout and threats to their mental health and wellbeing. This study has further enriched our understanding of personal growth and trauma in adverse work conditions by including an exploration of what sacrificial commitment adds to our understanding of physical and moral courage. Future provision of supports for nurses must be ongoing both during and after crisis events.

## Introduction

Long before the COVID-19 pandemic, the mental health and wellbeing of nurses has been a concern, and they were already overstretched, due to the nature of their work, and also due to austerity measures during the last financial crash [1, 2]. During the pandemic, and again, as a result of the 2021 cyber-attacks on the Health Service Executive (HSE) in Ireland, nurses once again found themselves facing severe adverse events. This has resulted in nurses, and other healthcare workers experiencing significant change and adaptation while navigating the post-pandemic landscape.

Younger nurses aged between 18 and 35 years, who were married or in relationships, with dependent children, were amongst the most exposed of all healthcare workers [3, 4]. Older nurses, the majority also female, suffered considerable health impacts [5]. Interestingly, a recent systematic review suggests that almost 32% of nurses intended to leave the profession during the COVID-19 pandemic [6].

Redeployed nurses were “the hardest hit” [7]. Their workloads scaled up significantly both at work and at home. Women, but particularly nurses, were expected to prop up our healthcare system while also largely holding down the family home. This caused them much distress, overwhelm, and at its extreme, burnout, and even suicide [8], but some also faced these challenges with resilience and courage.

Nurse ‘redeployment’ during the pandemic was when nurses were assigned without choice, from their own specialist clinical settings to alternate and often unfamiliar COVID units [9]. As a result, they felt forced into highly complex situations where they had to rapidly flex and adjust, due to unprecedented disruption to their working lives, resulting in them experiencing poorer physical and mental health outcomes [10]. It is important to note that not all nurses were redeployed, and those nurses got to carry as normal in their own clinical settings without any major disruption.

Women make up more than 85% of the global nursing workforce [11], and it is well documented that the pandemic exacerbated gender inequalities in women, which has introduced brand new challenges that are unprecedented to society at large [12].

According to the Women in Global Health Report 2023 [13], ‘ingrained inequalities and asymmetrical power relations, gendered organizational structures and norms’ are at play to shape the experiences of healthcare workers. The Nursing and Midwifery Board of Ireland (NMBI) published data on the State of the Register 2023 [14] reporting that 90% (71,456) of all nurses and midwives registered in Ireland were female.

As a result of the profound effect on a majority female workforce, this study aimed to examine female nurses’ resilience in acute hospitals in Ireland, during a period of adversity (COVID-19 pandemic), to explore it’s impact on their personal and professional identity, and their perception of meaningful supports to cope.

### Sacrifice & Emotional Response

One of the key burdens facing frontline nurses was the higher risk of contracting disease which contributed to their feelings of fear of being infected, or by infecting others, including their family members or friends which affected their emotional, psychological & physical wellbeing and their capacity to do their job [15, 16].

Another key burden that nurses faced was the existential threat of death and dying, which created a heightened state of alertness and resulted in sustained levels of hypervigilance [17, 18]. Because this extra burden caused so much distress and threat to nurses, they relied on potent war metaphors to help portray their trauma [18]. War metaphor themes coming from some of the research included ‘hard armour’, ‘temporary concentration camps’, and where nurses reported having ‘won this battle’ [18, 20].

A further pressure experienced by the nurses was how the public portrayed them as heroes. This hero label placed too much emphasis to perform at such a high standard that a common trend among nurses, and other healthcare workers (HCWs), was to push the hero label away to protect themselves professionally [21].

Despite being redeployed to the frontline without choice, the nurses’ profound sense of duty and professional responsibility enabled them to stay, commit and sacrifice themselves in a harsh climate, flanked by their peers [19]. Furthermore, the literature suggests that the exposure to morally injurious events has been one of the most prominent factors for nurses during this pandemic [22].

### Experience of and adaptation to adversity (resilience)

In facing this fear, and emotional turmoil, nurses found meaning and purpose in their new roles, resulting in increased levels of self-efficacy, thus learning to adapt well in the face of adversity [23]. Furthermore, emotional toughness, personal character strengths, and being able to detach, have all been identified as strategies used by nurses to build feelings of hope and self-efficacy, thus increasing resilience [24]. During the pandemic, resilience played a protective role where positive factors such as tenacity, strength, and optimism, were observed [24]. On the other hand, what has been shown to reduce nurses’ resilience is work conflict, poor organisational and social supports, feeling undervalued at work, exhaustion, moral injury, defective preparedness, poor communication, and constantly changing guidelines [15].

### Post Traumatic Growth

Nurses can experience post traumatic growth after traumatic events, and this higher level of functioning can coexist with the ongoing distress experienced during traumatic events [25]. For frontline workers and nurses, those with higher levels of perceived solidarity with social groups, were found to have a stronger sense of purpose and meaning in life, and pride in their work, resulting in growth [8], Because nurses are perceived to be caring and compassionate, practicing those virtues and knowing their own personal characteristics and strengths can help bring some meaning to their adversity as well as to build hope and self-efficacy to bring them some work life balance [20].

### Supports

Nurses coping mechanisms and social supports have been shown to strengthen their resilience when faced with adversity and crisis. Apart from Maslow’s fundamental hierarchy of needs, both intrinsic and extrinsic factors play a role in how nurses support themselves [26]. The importance of self-care has been shown to impact positively on all healthcare workers during the pandemic [15, 27]. Also, detaching from social media, news, and government briefings was a protective coping mechanism for nurses [21].

Nurses also relied on other self-coping mechanisms such as introspection, engaging with nature and virtues (practicing gratitude and having hope and wisdom) to get them through the crisis thus making them more resilient [28]. Notably, at times of severe adversity, some studies found a strong link between spirituality and improved health, such as the ability to forgive oneself [29].

Although in general, supports had a positive effect, there were certain times when supports were less helpful. For example, although nurses were brought closer by their shared experience, there were also times that relationships with their peers were strained, especially where workloads were reported to be unequal and where there was workplace incivility [27]. Another example was where nurses reported feeling touched at the acknowledgement and generosity from the public during the pandemic, which initially boosted morale and confidence, however, they described this relationship as a ‘double edged sword’ where most felt it was short-lived once the clapping and support from the public had waned [15, 16, 27]. Also, nurses who sought support from family, found that they were not able to relay to them what they were going through, and although they were ‘in the same boat’, or ‘in it together’, this did not resemble the collegiality and support they felt with other nurses [30].

Frontline clinical nurse managers also provided support to nurses, and other healthcare workers, both during and since the pandemic, in the form of workplace interventions. Some workplace interventions focused on having psychologically savvy, ‘on the ground’ clinical nurse managers who were adequately equipped and trained to support nurses in crisis [31]. Other studies that focus on nurses and other healthcare workers, indicate that experienced leaders need to have early detection strategies, to support and address any mental health risks. For example, Greenberg and colleagues recommend that managers need to: be empathic and listen out for staff that may be at risk; only offer low level psychological interventions early, followed by a clear pathway for staff to access professional help, such as trauma risk management and peer support if required [32].

Nurse leadership during the COVID-19 pandemic was reported to have been more meaningful where leaders were actually present, where they showed vulnerability and where they stepped up to acknowledge challenges, even if they didn’t have all the solutions [33]. In stark contrast to that, a UK study identified how little presence there was of senior nurse leaders on the frontline in the UK during the pandemic; they tended to focus more on policy and procedure over the power of compassionate leadership and emotional support for staff [23]. This further highlighted the importance of a visible presence of senior nurse leaders on the frontline, who prioritised ‘shared models of engagement [23].

Formal supports for nurses can also include evidence-based strategies or interventions. Clinical debriefing sessions provide a psychologically safe space for nurses and their co-workers to share their thoughts and experiences and to process emotion, enhance mental health, and enable them to appreciate each other’s contributions. However, it is now well accepted that debriefing should not take place during crisis events, not that it doesn’t work but that it can cause harm. Instead ‘psychological PPE’ (not debriefing) is recommended during a crisis event at work, where nurse leaders adopt a ‘nipping it in the bud’ approach, providing nurses with a focus on early ‘return to duty’ where at risk nurses are actively monitored in a ‘watch and wait approach [34].

It is important to highlight that despite the HSE’s significant investment in mentoring and coaching, there was no formal attempt to roll out supports through these existing services, either during or after the pandemic. This may have been beneficial in helping HCWs to heal, while also engaging in post-traumatic growth and adaptation.

Positive psychology, the fastest-growing discipline in applied psychology, is closely associated with post-traumatic growth, and is gaining considerable traction in healthcare worker wellbeing research [35]. A recent systematic review strongly indicated that positive psychology interventions (PPIs) show promise for improving the wellbeing of healthcare workers in crisis, albeit with more focus being placed on organisational-led as opposed to individual-led interventions [36].

Some qualitative studies have given insight into frontline healthcare workers’ experiences in the early stages of the pandemic. An interpretative phenomenological analysis (IPA) study explored UK healthcare practitioners personal experience and contextual factors and how they influenced their wellbeing in the early stages of the pandemic [16]. Whereas in another phenomenological study, the researcher gave insight into how doctors and nurses, working in China during the pandemic, ended up taking a lot of responsibility, thus explored their resilience, and the challenges they faced at work [20]. Some qualitative research from other countries, including the USA [33], also focused specifically on nurses’ experience of caring for their patients during the pandemic. However, understanding and insight is needed into Irish nurse’s experience, especially their resilience during adversity, that of the pandemic, and what was supportive for them. This is particularly important given how stretched Irish healthcare workers have been for so long, due to austerity, attrition rates, and the recent pandemic.

## Materials and methods

### Aims

The aim of this study was to examine nurses’ resilience in acute hospitals in Ireland, during a period of adversity (pandemic); to explore it’s impact on their personal and professional identity, and their perception of meaningful supports and coping.

### Design

This is a qualitative IPA study exploring the thoughts, feelings, and perceptions of the nurse participants. The researcher drew upon IPA to explore a more existentially informed study. The two key principles of phenomenological psychology fit well with this research study as it focuses on intentionality, or ‘what’ is being experienced, and how one’s biases add further constructs of meaning to the experience. Hermeneutics is the art of interpretation, and the second theoretical foundation of IPA where the researcher attempts to step into the shoes of the participants, though that may never be completely possible [37]. The third theoretical foundation, idiography, is the in-depth analysis of individual participants, and the examination of their perspectives in their own unique contexts [37] researcher actively engaged with the transcripts in analysing what the participants were saying, but also at how they were saying it, allowing the researcher to read between the lines thus developing a thorough understanding of the personal experiences of each participant.

### Study participants

Six (n = 6) female nurses between the ages of 21 and 65 took part in this study, the author’s master’s dissertation. All participants were redeployed from their existing roles to COVID wards in acute Irish hospitals, during the pandemic, from two provinces in Ireland. The nurses were recruited, under very difficult clinical circumstances, via convenience and snowball strategies, which represented a homogeneous sample of participants in keeping with the theoretical pillars of IPA [38]. When it comes to IPA, Reid and colleagues suggests that ‘less is more’ in its commitment to ideography, and that the examination of fewer participants in greater depth is far more valuable than the simple descriptive analysis commonly seen in grounded theory, thematic analysis, or indeed in weak IPA studies [39]. Smith and colleagues report that sample size depends on study context and that each study should be considered on a case-by-case basis. They suggest that between three and six participants for master’s level is sufficient [40].

Only female nurses who were redeployed to COVID units were included in the study. The rationale for this was that female nurses who were redeployed to COVID units, and who had significant caring commitments at home, were at greater risk of psychological distress and harm [41]. Consequently, male nurses and student nurses were excluded from the study. Nurses who were not redeployed or who did not work on COVID units were also excluded.

### Data collection

Recruitment for the study commenced on 8^th^ February 2021 and ended on 1st March 2021. Written communication took place via the researcher’s university email credentials and verbal correspondence took place via the researchers Microsoft Teams account. Following initial email contact, a short Microsoft Teams call took place to provide a brief outline of the study, obtain verbal agreement, form initial rapport, and address any queries. Participants also received information on the purpose and nature of the study, the aims and methodology, and information about the researcher. Following the calls, each participant was emailed a copy of the study information sheet and consent form as well as a copy of the interview schedule. Signed consent forms were returned to the researcher by email and stored securely on OneDrive, the university’s secure server. The interviews took place between 15^th^ March and 1^st^ May 2021, during the latter end of the third wave of the pandemic. Participants were briefed again at the start of each interview on the reasons for doing the research, and that the information they provided during the study would be kept confidential, and that they reserved the right withdraw from the study. The first author carried out the virtual semi structured interviews, lasting between 60 and 90 minutes (mean length = 75), using Microsoft Teams, due to lockdown protocols, and to protect the nurses. The recordings were split using Panopto software, video recordings deleted, and audio recordings were uploaded to OneDrive for short term storage to facilitate transcribing. Interview notes were also stored securely on OneDrive. Once transcribing in Microsoft Word had concluded, transcripts were forwarded to UCC via edugate’s authenticated Filesender services for long term storage on an encrypted hard drive, and all associated OneDrive files were subsequently deleted. Microsoft Excel was used at the analysis stage and for table formatting. The interview schedule followed a topic guide consisting of four main domains; experience of and adaptation to adversity; self & identity, supports, and checking in and debriefing. Semi-structured interviewing was chosen as it allowed a purposeful conversation; beginning with general questions, moving to more personal questions once rapport was established. It also highlighting the subjective experiences of the participants, while also taking into consideration the use of language, cultural norms, and social aspects of the data [42].

### Ethical considerations

Ethical approval (application MCP 812202005) was obtained on 16th December 2020, from the School of Applied Psychology Research Ethics Committee (APREC), at University College Cork (UCC). Participants were informed prior to the interviews, the reasons for doing the research, and that the information they provided during the study would be kept confidential, and that they reserved the right withdraw from the study.

They were informed that their names would be removed by being assigned numerical codes during the transcription and coding process, and by using pseudonyms during data analysis. The researcher also checked what supports participants had access to, and provided additional support where required, and given the stressful nature of the participants’ work, the researcher checked in with participants at the start and end of each interview.

### Data analysis

Interviews were transcribed verbatim by the main researcher (SC), and analysis took place by drawing upon the seven steps of IPA [40], to explore a more existentially informed study. The aim was to generate, categorize and organise potential themes from the data by reading and re-reading transcripts and listening to the audio recordings, where the researchers stepped inside the world of nurses working on COVID wards, to try and understand how they experienced it, and to examine how they made sense of it, as meaningful insights were crucial to the understanding, perspectives, and common experiences of each nurse [40]. Any queries from the findings were directed to the participants for clarification. Transcripts were analysed individually and were annotated by colour-coding and note taking to try and identify potential codes and themes described as descriptive, linguistic or conceptual concepts. The next step required the researcher to identify and label the emerging themes from the transcripts which represented the nature, quality and meaning of the nurses’ experiences, including themes such as ‘intense fear’ and ‘potent use of metaphors.’ Coding was cross checked for errors and omissions with the second researcher (AT) who was also the author’s dissertation supervisor. The final steps involved an attempt to introduce structure where the researcher listed themes, and formed clusters with shared meanings and references, and others that were characterized by hierarchical relationships with each another. The researcher moved back and forth between themes and original text where some changes to codes were made in this process. The final step involved producing a summary table with quotations

### Role of the researcher and study rigour

One of the researchers (SC) was a senior frontline nurse working in a critical COVID-19 role in the acute hospital setting. Because at times, she felt emotionally involved, and understood what her colleagues were going through, she maintained a journal and checked in with her supervisor AT, as a strategy for maintaining reflexivity, keeping track of reasoning, judgment, bias, and emotional reactions [43]. It should be noted that SC was analysing the “familiar” rather than the “unknown” and an advantage here was that she had an inherent feeling of what the participants were going through, and at times could recognise, and have the coaching competence to refer participants for external supports where required. The researcher SC implemented certain strategies to ensure quality, credibility, and trustworthiness during data collection and analysis which included an experienced researcher AT providing oversight and feedback on the interview protocol, analysis process and themes. Regarding transferability, within the write-up, the researcher SC provided adequate information for readers to evaluate relevancy, whilst also evoking a sense of shared experience. Further details can be viewed in supporting information.

## Results

Between March and May 2021, the first author interviewed six (n = 6) experienced nurse participants between the ages of 21 and 65 years. Five (n = 5) were Irish nurses and one (n = 1) was from outside Ireland but was working in Ireland for most of her nursing career. One participant was approaching retirement. Participant characteristics are summarised in Table 1. Three experiential themes were found in this study that illustrated the nurses journey from firstly needing to find ways to protect themselves from the sacrifices on the frontline, to then moving to fortify themselves as a collective, and finally through experiencing critical turning points and growth. A summary of superordinate, subordinate, sub-themes & quotations on nurses’ experiences on the frontline during the COVID-19 pandemic are listed in Table 2 and are further illustrated in the thematic mapping below (Fig 1).

**Fig 1.** Thematic map illustrating three major themes and subtheme.

**Table 1.** Participant characteristics.

**Table 2.** Summary table superordinate, subordinate and sub-themes & quotations.

### Protection of Sacrificial Self

Nurses found a need to protect themselves emotionally and mentally from the intense psychological overwhelm they experienced, which was triggered by the sacrifices they made on the frontline.

The emotional intensity arose from the nurses coming face-to-face with an existential threat, and the moral distress they experienced by being deployed to an unknown territory. Their fear was potent, especially the fear of infection and a fear of death and dying, for themselves, but also for their loved ones and patients.

> Well, you walk into a ward, you might not have been in there for three hours, you don’t know what you’re going to get … there are bodies going out every single day. There are just all these people crowding in. It’s like “Mash” … coming in on trollies, barely able to breathe, frightened, anxious. I mean, it was a war zone (Una)

Although they were redeployed to the frontline, the nurses showed a strong willingness to turn up and serve every day. This stemmed from a moral duty where their values got called into question in situations where they felt dangerously ill-equipped to cope.

> I wasn’t fighting the fact that I needed to go in and do this… I knew it was the right thing to do, so there was no doubt in my mind even if I got COVID. (Una)

Concerns about the nursing care deficits, meant that some even volunteered extra time with their patients to address this:

> I sat with quite a lot of patients … so they wouldn’t be on their own, which I wouldn’t have done before, but I made an effort … I’d stay on after shift… and just give them that little bit extra attention and care, just because they weren’t going to get it, when, when they did die (Emma)

Claire portrayed a compassionate picture of how devastating it was for her, not to be able to provide dignified care at end-of-life. She suffered extreme guilt and despair at being forced to pack her patients’ bodies into cadaver bags in their hospital gowns, which deeply wounded her, as it went against her core values:

> I don’t want people to go in double black bags … I feel like the dignity of the person at the time of their death, or even after they died, I’m not a grocery material to go in a plastic bag … I don’t want to pack anymore bodies; I don’t want to do that (Claire)

All the nurses felt betrayed and lonely where they were left to face high risk and ethical dilemmas. Three of the nurses, in more critical frontline roles, felt forced into choosing which patients to save over others, and this came with huge emotional cost. “I took on everything, and if someone was deteriorating, it was because I wasn’t doing a good enough job or if I had missed something…I felt like it would have been my fault” (Emma)

The nurses identified ways of protecting themselves emotionally to help maintain their focus at work. Some put their own emotions on hold whereas others just about ‘held it together,’ and all thought they would fall apart if they began processing their feelings too soon:

> I can’t let myself fully process the past year, because I don’t know if I’m gonna fall apart or not, … am I gonna hate my job and never want to go back…if we start kind of questioning ourselves, am I OK to do this (Trish)

Another protective mechanism was that most of them purposely detached from their chaotic surroundings. Ruth recalled how vital it was for her to “get off the merry-go-round” to recharge before getting back on. Three of the nurses detached by mentally hunkering down to unshackle themselves from the enormous mental strain. Once hunkered down, they entered into a purposeful flow state, with a heightened focus on critical tasks. This induced flow-state was protective in nature as it helped them to focus on the present moment, shelve their distractions, which helped sustain their energy levels:

> My mantra was “one day at a time, one task at a time” and the only way I could get through my day, and if I lifted my head up to think of all the things that I had to do and all the responsibilities I had, I would have panicked and not being able to do anything, so I had to just break it down. “I’m here, I have to do this, no one else is going to do this, I have to just go in and do my best, there is nobody else” So, you need to just concentrate and try and get through the simple tasks that you’re doing and then you go on to the next task and … my main aim was not to kill anybody (Una)

Another way they protected themselves was by the potent use of metaphors which helped them to make sense of the complex situations they found themselves in. Quite often they likened traumatic events to war where they endured “the scars” of battle by “working in the field.” Trish gives us a sense of how courageous and fearless she felt going into battle with “all guns blazing” flanked by her peers.

### The Fortifying Effect of Us

As a result of feeling isolated, unsupported, and left out in an unrelenting climate for far too long, the nurses turned to each other and began to develop a strong solidarity or “us”, which helped them to bolster each other and build self-esteem against a growing perception of “them and us”.

Frontline nurses were left feeling unsupported by a public who once portrayed them as heroes, but now blamed them for spreading infection in hospitals. They also felt more isolated as they were the ones left holding the burden of responsibility, while others flouted guidelines or were spreading hate speech and vaccine misinformation on social media. This frustrated and angered Claire who wanted others to witness what was really happening behind hospital doors:

> Am I allowed to say that I would love to go slap people who say there is no such thing as COVID? … because they haven’t seen what we have seen in that place, they were not there holding the hands of a person that was dying (Claire)

As a result of feeling isolated, all the nurses, over time, lost trust, and confidence in how the public supported them, and what began to emerge was a growing divide between “us”, the nurses, and “them”, the general public and media. This was further compounded by feelings of abandonment at work, because they were excluded from key decision-making, and were not being adequately supported. Trish described how the lack of presence of meaningful leadership and supports, led to feelings of extreme hurt and loneliness, “to even see a face …, would have meant a lot to us…one visit … to the unit … it really made us feel aware of where we are in the pecking order” (Trish)

The nurses felt even more isolated as they perceived their redeployment as unfair, and resented being called to the frontline over and over again by senior management, when compared with other nursing colleagues who were never once redeployed.

> I think a lot of us just felt like it was a kick in the teeth…we really just felt like a number…you know, redeployment was part of the gig, so “what do they have to complain about?” so yeah, it was just disheartening (Emma)

The workload disparity and constant presence on the front line, was emotionally draining, placing an even bigger wedge between “us” the nurses, and “them”, other healthcare professionals.

> It’s more difficult because we’re with the patient all day, I mean the rest of them will do their treatment and … go, the doctor will come in and do their treatment and go, the dietitian will come in and do her treatment and go, you know? So, we were the ones that were there all the time, yes, the constancy of it (Denise)

In addition, the nurses were unable to lean fully on family and friends for support because they felt that they couldn’t possibly have understood what they were going through. Although they weathered the same storm, they were not all in the same boat as the nurses. Nevertheless, both Una and Emma at times, felt their partners were “key” to getting them through emotionally:

My poor boyfriend then got it all … if I was quiet or upset … he knew when to push and pull back with things, which was great (Emma)

At a certain point, things got so gruelling for the nurses that they began to question why they ever chose nursing as a career. Denise, who felt disrespected and undervalued, said, “I don’t love nursing as much as I used to, and I would like…to maybe leave and pursue something else … I don’t feel that my service was valued” (Denise)

What began to emerge for the nurses was a sense of power in numbers and solidarity. This is how they supported and bolstered themselves in a harsh climate, and began to build strength and confidence, feeling stronger against “them”, and stronger against the world:

> But from the lessons learned, I would (pause)… look after myself and look after my colleagues, that’s all I can do … because that was one thing that was poorly done, nobody ever had come back to us even asking us how we felt (Claire)

The sense of “us” gave hope at a time when commitment was needed. Claire described how the strong bonds she had developed with her colleagues served as a lifeline to her when she needed support most, “We kind of like poured our hearts out and we were able to…vent our feelings with each other, to share our emotions” (Claire)

This solidarity delivered a huge amount of renewed self-assurance which helped protect and re-establish their self-esteem. Furthermore, the nurses also began to feel the positive relational effects of bonding together, “you’re just tighter with your colleagues because you had to get through this together” (Una)

At times, some of the frontline clinical nurse managers (CNM) also became a part of us:

> He was just one of those grounding people that if you saw him being calm and collected, you knew you could be as well…He knew our abilities, he knew what we could and couldn’t do, he knew when to check in on us … he was a great voice for us (Emma)

Another CNM provided a positive road map forward which helped build inspiration and enthusiasm within the nursing team.

> The CNM came up with an idea called the positivity board … It brought smiles to our faces, and I remember one of them writing “thank God the numbers are coming down”, or … “I did not have to put anyone into a black bag today” (Clare)

Tributes at work were a stark reminder of how far the nurses had come and how lucky they felt to come through it:

There … were pictures of us, socially distanced together; so those pictures went up on the positivity board, something to look back and say “OK, fine we have survived this (Claire)

All the nurses felt a conjoined sense of meaning and purpose with their nursing colleagues around the world which gave them immense strength:

> I think there’s some kind of comfort there as well to know that it’s happened all around the world and … most nurses have gone through it all together, so there is some kind of unity yeah, and like solidarity (Emma)

### Critical Turning Points and Growth

Being away from the frontline allowed them to look back and accept their feelings of fear, anger and overwhelm:

> So, anger was the first thing, like there was no reason to be angry with everyone, I was angry within myself … I wasn’t able to get on top of doing my things, which I would have normally done on a day-to-day basis looking after my patients (Claire)

Engaging in positive self-talk was another turning point. Positive self-talk enabled them to feel less of a victim, reaffirm their contributions and to feel more in control, “I just had to talk to myself and tell myself I could do it and just get on with it and stay calm and stay focused” (Una)

Feeling better able to cope was another turning point. Claire felt if there was another COVID surge, she would be willing and happy to do it all again, having earlier felt she could never go through it again, “I don’t want another wave coming, but if it does come to it, I would be willingly and happy going there” (Claire)

Another critical turning point was the ability to visualise to feel hopeful. For example, Denise, was reminded in her own drawings that all waves eventually diminish, and that the safety of her lifejacket would carry her through:

> I had one particularly bad shift … I remember getting up the next day … and painting surfing waves … and then there’s this tiny little figure in a life jacket down at the bottom and that’s me. It ended up that it really helped you know, because … I’m OK, I will survive this so (Denise)

Similarly, Una expressed deep gratitude towards a child who had drawn a picture of a nurse superhero, and this struck a chord with her, giving her strength to keep going, “Thank you, our heroes, and I saw it and I just burst out crying like you know, that child, he doesn’t know that, but…it did make a massive difference” (Una)

Another critical turning point was being able to reconcile with self:

> I had feelings that the care delivered to the patients was sub optimal definitely, some of the time, not all of the time, and I had to reconcile that with myself with ‘I did my best, I did my very best’ (Ruth)

A renewed meaning and purpose in their work and a strong sense of team spirit and pride was another turning point, “I’m proud of myself for going in and having put my shoulder to the wheel and, you know, pushed with the rest of my colleagues … I didn’t run away” (Una)

Most of the nurses now felt more confident and vocal. For example, Emma’s expert knowledge made her feel “more confident and definitely more resilient”, with a strong desire to become a future nurse leader:

> Well as a nurse, I think I’ve found my voice: if there is a sign of deterioration or anything … I feel more confident in myself. Definitely more confident in my ability and my experience and my knowledge (Emma)

What was strong for all the nurses was the pride and gratitude they felt for the work they had done:

> I might have not done everything, but I was there for someone, and I’ve helped out, I have touched someone’s life. I was able to offer that service, and I feel so good about it myself, that I have chosen this profession (Claire)

All of the nurses begun to take great comfort in ordinary things, such as returning to their families after feeling so isolated on the frontline. They also felt a deep sense of “appreciation” for just being alive and connected with nature:

> The sea is very calming, it’s just like meditation, it’s full of life, it’s so positive, … it’s the simple things that I enjoy …, baking in the kitchen, family and friends, a warm fire in the evening, a home cooked meal, barbecues in the garden, dogs, animals, cats (Una)

The interviews for this study fostered new meaning and purpose for the nurses and helped them to make sense of their journeys. This brought about a renewed faith in peoples’ desire to help and listen to their stories, “You touched on every aspect of it and it’s great, like, I feel this itself is a debriefing session … just kind of talk about it and thank you, thank you for doing such a study” (Claire)

## Discussion

Three key experiential themes were found that represented various stages of a journey taken by the nurses: first was the need to protect themselves emotionally and mentally from the intense psychological overwhelm they had experienced, having sacrificed themselves on the frontline; the second theme arose due to the nurses feeling isolated, and left out in a harsh pandemic climate for so long that they turned to each other for solidarity to bolster each other, and build self-esteem. What began to emerge was a sense of power in numbers which made the nurses stronger with each other and against the world; Finally, all of the nurses began to experience some critical turning points and growth.

### Protection of Sacrificial Self

The current IPA study gives insight into how nurses emotionally and mentally protect themselves against the sacrificial commitment they made, and moral injury experienced, due to having to sacrifice their own high standards, while facing intense fear, psychological overwhelm, and prolonged existential threat posed by the pandemic. The sacrificial commitment stemmed in part from the nurse’s strong willingness to turn up every day, despite the unreasonable challenges and mandatory redeployment.

Gaining insight into how the nurses protected themselves is important given that studies have found that nurses experience fear when at risk of contracting disease, thus impacting upon their emotional, physical and mental wellbeing as well as their capacity to do their job [15]. Some studies have found that the threat of death and dying creates a damaging heightened sense of alertness, and hypervigilance [16], and other studies report injury, and exposure because of upholding a strong moral, professional, and spiritual work ethic [17].

Some of the key ways the nurses protected themselves at work in the current study involved emotionally detaching, hunkering down, shielding themselves and making use of metaphors. Notably, some of them hunkered down to intentionally place themselves in a flow-type state, allowing them to focus on the present moment and distracting themselves from the surrounding chaos. This helped them to ‘hold it together’ and put their emotions on hold, so that they could continue to focus on the tasks at hand. Remarkably, only one study described a similar experience where ICU nurses were “pushing through long temporal episodes of work described as being in a “bubble” [44]. Theoretically, this study also contributes to the understanding of a hunkering down flow-type state, which may be an optimal state that can be achieved when under pressure, thus, adding further insight into Csikszentmihalyi’s definition of flow, which requires one to enter an optimal psychological state [45].

Protection of self was strong with the nurses in the current study who shielded themselves from risk in various ways. For example, they engaged in positive self-talk and mindful mantras during work to be able to sustain and maintain their hunkering down type flow state. This fits with some literature which found that positive self-talk can increase work efficiencies whilst also reducing stress levels [46]. Also, the nurses in the current study shielded themselves from risk by detaching from work to recharge, getting enough rest, and staying in touch with family and friends. They also reduced their interactions with social media and news reports which is a common theme found in the nursing literature [21, 27].

Another interesting way to protect themselves emotionally was by use of metaphors in an effort to make sense of, and move forward from, the emotional trauma they experienced. The richness of the metaphorical language used by the nurses in the current study, was also evident in other nursing literature [18–20]. However, in contrast to the wider literature, the nurses in the current study also used metaphors to highlight positive emotions such as courageousness, fearlessness and pride.

### The Fortifying Effect of Us

Another key experiential theme in this study was the fortifying effect they gained from being an “us” against “them”; them being the general public, politicians, and at times their own family, and senior managers. Nurses turned towards each other over time to look for back-up, and reassurance, when public opinion and support, had moved from being portrayed as heroes to waning support. Nurses found themselves mentally distancing themselves from the burden of the hero label in order to protect themselves. The above findings are consistent with some studies [21, 44], where the hero status placed additional pressure on nurses to work even harder.

Also, the nurses turned to each other, to find strength and support as they often felt alone and isolated from their own family members, who didn’t understand the distressing nature of their experiences with death and suffering. Although the nurses felt they were weathering the same storm, they were not in the same boat. Not surprisingly this was broadly consistent with the wider literature, such as where nurses found it hard to relate their feelings and experiences to their families as they just didn’t understand what they were going through [30].

Another strengthening of a sense of us came as they felt solidarity together as a group who were experiencing strong feelings of exploitation and abandonment at work. They felt excluded from important decisions, and most were redeployed without choice, irrespective of their qualifications, personal strengths, and experiences. Unfairness was also felt, because some nursing colleagues were never once called upon by management for redeployment. This created resentment and unnecessary strain on relationships between the nurses (us), and some of their colleagues (them), as well as between nurses and senior hospital managers. This fits with what Billings found among redeployed staff, including nurses, who held resentment against others who were perceived to be doing less work [27]. Furthermore, as a collective, some of the nurses started to question why they ever chose nursing as a career and some contemplated leaving the profession altogether.

What emerged from the above findings were feelings of isolation, aloneness and largely feeling unsupported in a challenging landscape. Over time, the nurses began to build alliances with each other (“us”) at a time when they needed to bolster their self-esteem and build resilience against ‘them’ and the world. This helped re-establish the self-esteem for which nurses were accustomed in their roles, and it gave them a renewed sense of hope at a time when huge commitment was required from them. This solidarity fits well with another study [44] which reiterates the importance of supporting one another to help cope with the situation.

At times other staff felt part of this collective ‘us’, such as the frontline clinical nurse managers (CNMs), who at times brought further support. This helped build some inspiration and enthusiasm on the ground and can be seen in the wider literature where stronger collaboration on the ground helped improve nurse wellbeing [44]. This fits with the findings in one thematic study [47], where nurses experienced a stronger sense of ‘them and us’ with senior managers, where a lack of their presence and support was felt.

### Critical Turning Points and Growth

The third experiential theme was that the nurse experienced critical turning points and growth as they wagered through the emotional turbulence of working during the pandemic. It has been strongly suggested that when nurses fail to do for their patients what they know deep down is good for them, their own moral integrity is at risk, and they end up with moral injury [48]. Not being able to alleviate pain and suffering or provide and care for critically ill patients during COVID has been a prevailing feeling among nurses in the current study, and moving beyond this phase was critical to their survival.

Key critical turning points for the nurses in this study included when they were able to step back and accept their feelings; enabling them to feel more hopeful or better able to cope, and being able to reconcile, or find purpose and meaning for themselves. This is widely consistent with older psychological constructs such as finding benefit [49] and enabling growth [21]. In the same way that grief and recovery do not follow an orderly process [50], the nurses began experiencing pivots and positive changes thus allowing them to regain some sense of self control in their lives. For example, Ruth recalled special moments with her patients in their last days before death and Una was deeply moved as she felt valued and a sense of pride after recalled children’s drawing of nurses in capes. Both examples show the importance of emotional connectedness with nurse and patient. Empirical research has long identified emotional connectedness as a key coping resource for nurses [51].

Nurses in the current study conveyed a strong sense of duty toward their patients arising from a deep sense of needing to care [19, 20]. This permitted them to feel immensely proud of themselves, and gave them a sense of purpose and meaning in their lives [8]. This was echoed widely in the literature where nurses were reported to having felt strengthened through adversity thus impacting positively upon patients’ lives [52]. Moreover, Claire was able to rekindle her strong passion for nursing having lost it during her time on the front line. Harmonious passion is known to contribute to sustained psychological well-being, thus preventing negative affect [53], and the strong positive effect of believing in a higher power in times of crisis [29], may have contributed to Claire’s growth. Some of the nurses said they now felt more expert in their roles. They felt listened to and respected, particularly by their medical colleagues, and this meant a lot as they felt more empowered to voice their opinions. This is consistent across the wider literature where the new roles nurses assumed during the pandemic provided significant reward [8].

The meaning of life is a fundamental factor of human existence, relating to the enormous existential power a person has to face in everyday challenges [54]. Denise expressed herself through art, Una and Emma made a grounding connection with nature and the outdoors. Others simply experienced a deep sense of gratitude and appreciation for getting through this and for life itself.

In terms of future research, it will be important in the coming years to track frontline nurses over time to see how they become resilient and overcome negative psychological effects or burnout. Future research could explore nurses spirituality to see if it extends their sacrificial duty, and to see if this better enables them to cope. In terms of positive organisational scholarship, it would also be interesting to know more about how nurses can purposefully enter a flow state or excelling zone, to enable them to detach and focus on the present moment to enhance their role. It may also be important to explore if future crises would merit redeploying nurses to areas that align with their core strengths and professional experience before being redeployed. Finally, given how cleverly nurses used time, space, and the extensive use of metaphors, a study examining the benefits of metaphor coaching for frontline nurses may be useful.

### Strengths and weaknesses

It could be argued that there are similar studies in the literature [55–58], however, those studies used more conventional approaches such as content analysis and thematic analysis, producing more generalised findings, and not interpretative phenomenological analysis, which involves a much closer examination of the experiences and meaning of the subject matter.

Moreover, one of the main strengths of this study was its capacity to give voice to frontline nurses who worked in Irish hospitals during the pandemic, whereas most of the studies outlined above took place in other countries. At the time of writing, this study was the first to examine the thoughts, feelings, perceptions and resilience of nurses working on the frontline, through an interpretative phenomenological lense.

Another key strength is that the researchers have suggested a more streamlined definition for ‘nurse sacrificial commitment’. One of the most powerful findings was how the nurses purposefully entering into a flow-type state or excelling zone to detach from the surrounding chaos, which may enhance our knowledge on the psychological concept of “flow” in nurses during crisis.

One limitation with this study was that findings were drawn from six participants (n = 6) living and working in two provinces in Ireland. Although this offers a rich perspective from an Irish context, the researcher would have valued a wider scope of study participants but was unable to do so due to the overwhelming working conditions and time constraints on the nurses during the main COVID surge in 2021.

Another limitation was that male nurses working on COVID wards were not included in this study. Future research might include research on male nurses, or indeed a study comparing the effects of the pandemic on both female and male nurses. Some would argue that collecting qualitative data online can lead to limitations, however this was not our experience. In fact, participants reported that the online interviews were far more favourable than in-person interviews given the pandemic circumstances, and they felt much more at ease and protected from potential infection. All of the nurses valued deeply the online interviews as they felt they were the only meaningful form of support and debriefing they had received.

## Conclusion

Each of the nurses provided their own journeys about how they protected themselves given the sacrifices they were making, whilst moving to a collective sense of us, to gain back strength and resilience, and over time how they experienced critical turning points and growth. The analysis has given a deeper insight into the journey the nurses took to overcome the adversity they experienced in the Irish healthcare system. Moreover, they also learned to protect themselves, to bring balance to the sacrificial commitment they made as frontline staff, and to turn towards each other, creating solidarity, or an ‘us’, as well as bringing support, and building self-belief, which led to some critical turning points and growth.

The study also exhibits how strongly the nurses engaged in various self-coping strategies, most notably by the extraordinary use of metaphors to “grasp the meaning of highly burdensome experiences” [48], but also, by adapting daily work practices by purposefully entering into a flow-like state or excelling zone to detach from the surrounding chaos.

Provision of supports for nurses, and indeed all healthcare workers, must be ongoing both during and well after crisis events. Although there was strong evidence of peer-to-peer support, and support provided by a small number of frontline clinical nurse managers, none of the nurses in the current study had received any meaningful psychological supports from their organisations either during or in the immediate aftermath of the pandemic, despite their enormous sacrifice. Four years on, nurses in Ireland are still experiencing deeply, the negative mental health and wellbeing impacts of the COVID-19 pandemic. We must now direct our efforts towards how nursing professionals will be able to care for their patients, as well as being cared for themselves [59] as we face the next crisis which is looming on the horizon.

## Data Availability

All data can be shared publicly apart from anonymised individual interview transcripts because ethically there is a risk that this would compromise participant anonymity and confidentiality. Raw format (verbatim transcripts) contains both potentially identifying and sensitive participant information (e.g., detailed discussions, mental health challenges, and potential participant geographical locations). However, cleaned data (semi-structured interview guide, sample IPA colour-coded theme generation table, copy of the consolidated criteria for reporting qualitative research (COREQ), copy of critical appraisal skills programme (CASP) checklist, can be accessed at https://doi.org/10.5281/zenodo.13882448 for this research article. Data are available from School of Applied Psychology Research Ethics Committee (APREC), at University College Cork (contact via) for researchers who meet the criteria for access to confidential data.

https://zenodo.org/records/14174070

## Acknowledgements

To my six participants, you gave up so much to care for your patients and you became their family in their time of need. Thank you sincerely for your unrelenting sacrifice, and for your part in this research paper. Your stories have now been heard. Thank you also to my co-author and supervisor, Dr Anna Trace, for her support and guidance throughout the entire project.

## Copyright / Creative Commons

This is an open access article distributed under the terms of the Creative Commons Attribution 4.0 International, which permits unrestricted use, distribution, and reproduction in any medium, provided the original author and source are credited.

## Data Availability Statement

All data can be shared publicly apart from anonymised individual interview transcripts because ethically there is a risk that this would compromise participant anonymity and confidentiality. Raw format (verbatim transcripts) contains both potentially identifying and sensitive participant information (e.g., detailed discussions, mental health challenges, and potential participant geographical locations). However, cleaned data (semi-structured interview guide, sample IPA colour-coded theme generation table, copy of the consolidated criteria for reporting qualitative research (COREQ), copy of critical appraisal skills programme (CASP) checklist, can be accessed at https://zenodo.org/records/14174070 for this research article.

Data are available from School of Applied Psychology Research Ethics Committee (APREC), at University College Cork (contact via) for researchers who meet the criteria for access to confidential data.

**Citation:** Creedon S, Anna T. From protection of sacrificial self to critical turning points and growth: Nurses’ experiences on the frontline during the COVID-19 pandemic. Zenodo; 2024.

## Supporting Information

**S1 Table. Participant characteristics**

**S2 Table. Summary table superordinate, subordinate and sub-themes & quotations**

**S1 Fig.**
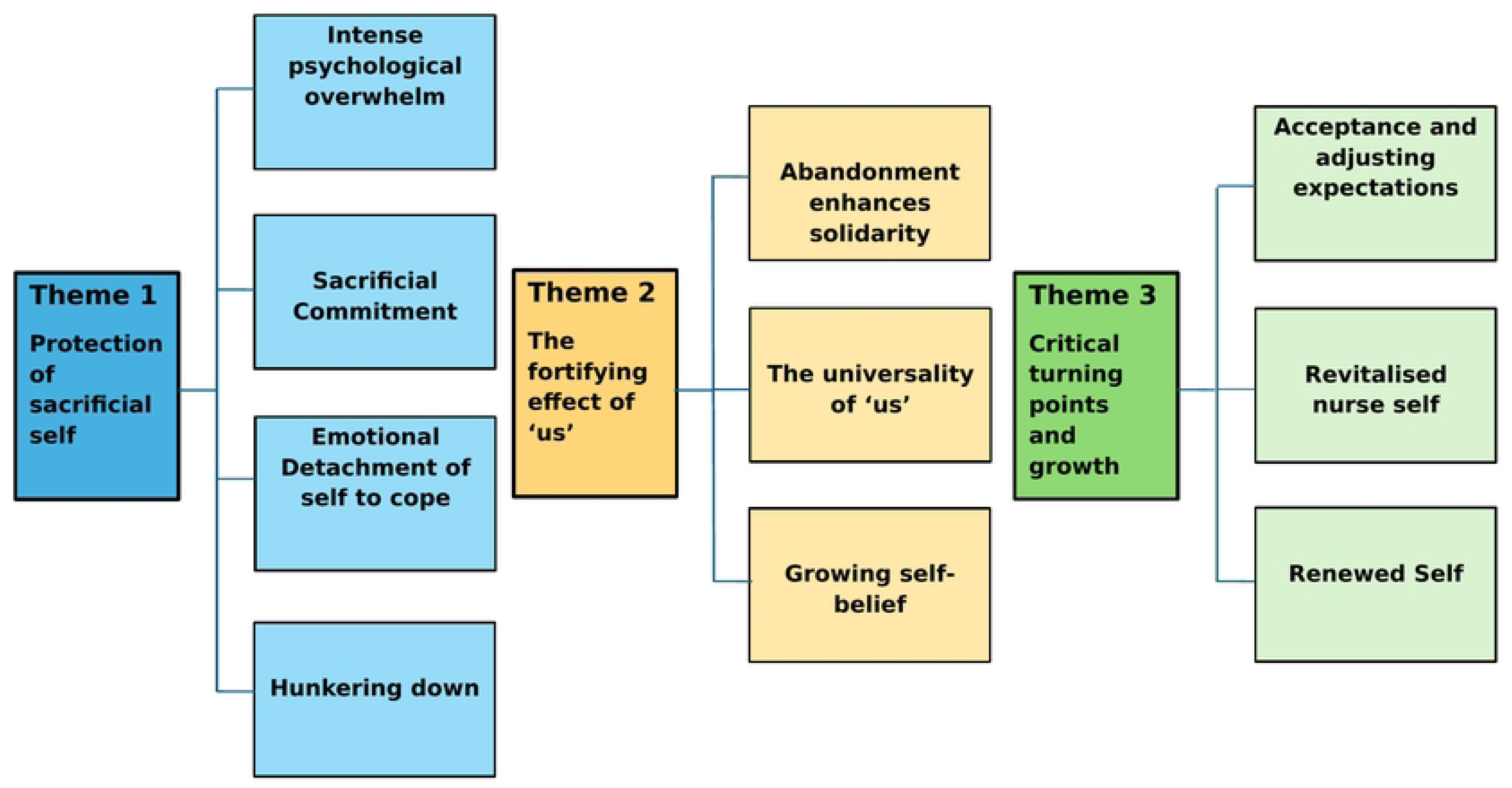
Thematic map illustrating three major themes and subtheme.

